# Taking injectable PrEP to scale: Optimising the value of lenacapavir for South Africa’s HIV response

**DOI:** 10.64898/2025.12.14.25342211

**Authors:** Lise Jamieson, Leigh F. Johnson, Jeffrey W. Imai-Eaton, Hasina Subedar, Linda-Gail Bekker, Gesine Meyer-Rath

## Abstract

**Background:** South Africa accounts for 20% of the global HIV infections and has one of the highest HIV incidence rates in the world. Six-monthly injectable lenacapavir (LEN) for HIV pre-exposure prophylaxis (PrEP) has superior efficacy to oral tenofovir disoproxil fumarate/emtricitabine (TDF/FTC), and similar efficacy to 2-monthly injectable cabotegravir (CAB). With LEN’s recent regulatory approval and the newly negotiated generic price of $40 per person per year, South Africa faces critical implementation decisions amid constrained domestic resources and reduced international funding. We evaluated the epidemiological impact on HIV infections and life years lost, cost-effectiveness, and optimal populations for LEN roll-out in South Africa.

**Methods and Findings:** Using Thembisa v4.8, a deterministic compartmental HIV transmission model of the South African HIV epidemic, we simulated the impact of LEN scale-up, expanded oral TDF/FTC, and CAB scale-up, compared to a baseline of current TDF/FTC provision over 20 years (2026-2045). We scaled PrEP among adolescent girls and young women (AGYW), female sex workers (FSW), pregnant and breastfeeding women (PBFW), men who have sex with men (MSM), and heterosexual men. For TDF/FTC scale-up, we doubled baseline initiation rates. For LEN and CAB, conservative and optimistic scenarios assumed initiation rates similar to or double those under TDF/FTC scale-up, respectively. Duration of use varied by subpopulation under TDF/FTC (3-6 months), conservative (LEN: 6-12 months, CAB: 4-8 months), and optimistic (LEN: 12-24 months, CAB: 8-16 months). We modelled strategies to maximise the impact of ~500,000 LEN doses currently allocated for 2026-2027, or large-scale roll-out. Costs are presented from the South African government’s perspective in undiscounted 2025 USD.

Providing LEN to 1.7-2.9 million South Africans per year averted 19-31% of infections and saved 3-5% of life years anticipated to be lost to HIV, reaching incidence <0.1% in 2039-2043, 10-14 years earlier than baseline. Estimating the number of individuals needed to initiate each PrEP type to avert one HIV infection, LEN required 35-65 initiations/infection averted, compared to CAB (45-125 initiations/infection averted) and TDF/FTC (280 initiations/infection averted). TDF/FTC and conservative LEN scale-up increased HIV programme costs by 3%; however, LEN was more cost-effective, costing $2,301-$3,567/life year saved (LYS) versus $8,143/LYS (TDF/FTC scale-up) and $11,114-$16,118/LYS (CAB). Prioritising PBFW, MSM, and FSW for the initial allocation maximized infections averted. Large-scale roll-out strategies prioritizing FSW and MSM were most cost-effective ($276-$958/LYS). Limitations to this study include uncertainty in achieving modelled uptake and assumptions of risk-differentiated uptake of long-acting products, and limited data on real-world implementation costs.

**Interpretation:** Delivering LEN to persons with elevated HIV risk in South Africa is more cost-effective than existing PrEP options and can speed up HIV incidence reduction. Prioritising uptake among groups at highest risk is essential to maximise impact and cost-effectiveness.

## Introduction

South Africa is home to the largest population with HIV, with an estimated 8 million people living with HIV (PLHIV), and, despite a 53% reduction in new infections since 2010, remains the country with the second highest rate of HIV acquisition worldwide.^1,2^ Lenacapavir (LEN), a 6-monthly injectable antiretroviral drug, for HIV as pre-exposure prophylaxis (PrEP) has demonstrated superior efficacy to oral PrEP with tenofovir diphosphate/emtricitabine (TDF/FTC) in the PURPOSE-1 AND PURPOSE-2 randomized clinical trials, both of which were unblinded early due to the extremely high efficacy of LEN.^3,4^ LEN is promising for HIV prevention, particularly in South Africa, which leads the world in oral TDF/FTC uptake, accounting for 20% (roughly ~600,000) of annual PrEP initiations.

LEN was recently approved by the South African Health Products Regulatory Authority for use as PrEP.^5^ To support rapid initial LEN implementation in South Africa, the Global Fund has reallocated $29 million of existing funding to provide approximately 500,000 person-years on LEN in 2026 and 2027 at a subsidised drug price to the South African HIV programme of $60 per person per year (PPPY).^6^ Beyond this, affordable access could increase through generic manufacturing: Six generic manufacturers have received voluntary licenses from the originator company to produce product for selected low- and middle-income markets, including South Africa, and two have committed to an introductory price of $40 PPPY (and $17 for a one-time oral loading dose at initiation) on expectation of sufficient market volume, a similar annual commodity price to oral TDF/TFC.^7^ Production cost estimates suggest eventual generic manufacturing price could be further lowered to around $25 PPPY for a market demand of 5-10 million people globally.^8,9^

However, strategic decisions for and success of LEN implementation will depend on several factors, including the pace of manufacturing, supply chain logistics, and creation of sufficient demand among and effective delivery models for those populations at highest risk of acquiring HIV infection who stand to benefit the most.^10^ Additionally, the readiness of public-sector facilities, including provider training, digital system adaptations, and capacity to manage injectable PrEP at scale, will directly affect achievable uptake. South Africa largely funds its HIV programme from domestic resources, making considerations of cost-effectiveness and affordability paramount. LEN’s recently announced $40 PPPY generic price and long-acting dosing may make it not only a cost-effective option within the existing PrEP landscape, but potentially a competitive investment compared to other HIV interventions, some of which may require substantially higher costs or coverage to achieve comparable impact.

Recent studies modelling the potential impact of LEN have shown that it could substantially reduce HIV incidence in sub-Saharan Africa. An analysis covering Eastern and Southern Africa—including South Africa, Zimbabwe and western Kenya—projected that LEN could avert 12%-18% of infections, over 10 years, at 1.6%-4.0% coverage, with higher coverage (3.2-8.1%) resulting in higher impacts (21%-33% of infections averted).^11^ In South Africa, a non-prioritized rollout strategy, providing 25 million person years of PrEP over a 35 years, was estimated to avert 4.4% of infections, while a risk-prioritized approach, which allocated to female sex workers first, averted three to five times more infections.^12^ Both studies generated threshold estimates for a LEN price that would render it cost-effective under a cost-effectiveness threshold of US$500 per disability-adjusted life-year. Results varied by epidemic setting according to underlying incidence, risk prioritization, and coverage scenario: Kaftan *et al*. estimated a range between $27-$106 PPPY depending on risk prioritisation, while Wu *et al*. estimated $177-$213 PPPY.^11,12^

To inform the prioritization of the planned roll-out of LEN in the South Africa, we conducted three analyses: (1) we estimated the cost and cost-effectiveness of a large-scale roll-out of LEN at the generic price of $40 PPPY, compared to CAB and oral TDF/FTC scale-up, and produced budget estimates for the South African government; (2) we compared both LEN and CAB roll-out to scaling up other existing HIV interventions, including testing, prevention and treatment; and (3) we modelled several roll-out strategies to different subpopulations to optimise the epidemiologic impact of the initial 2-year LEN allocation (~500,000 person-years annually in 2026 and 2027) (Phase 1) and the planned large-scale roll-out from 2028 onwards (Phase 2).

## Methods

### Epidemiological model

We used Thembisa (version 4.8), a deterministic compartmental HIV transmission model of the South African HIV epidemic.^13^ The model population is stratified by age, sex, sexual experience, sexual behaviour, marital status, HIV testing history and male circumcision status. The sexually experienced population is divided into two broad sexual risk groups: ‘high-risk’ (people who engage in concurrent partnerships and/or commercial sex) and ‘low-risk’; female sex workers (FSWs) are modelled as a subset of high-risk unmarried women, and men who have sex with men (MSM) are modelled as subsets of the unmarried low-risk and high-risk groups. PrEP uptake rates vary by age, sex and sexual risk group. Regarding sexual risk, the model assumes that PrEP uptake is proportional to the product of the individual’s annual number of partners and the HIV prevalence in their partners, resulting in higher uptake of PrEP of higher risk individuals. The model assumes that PrEP users (all modalities) have a 10% lower rate of condom use than individuals of the same age, sex and risk who are not using PrEP. Oral TDF/FTC effectiveness incorporates both efficacy and adherence, and was assumed to be 85% for MSM, and 65% for FSW, other women and heterosexual men.^14–16^ LEN effectiveness was assumed to be 99% for all populations modelled, based on data from the PURPOSE trials.^3,4^ CAB effectiveness was assumed to be 95% across all populations.^17,18^ For both LEN and CAB, we assumed that a pharmacokinetic tail protection is present beyond the respective 6- and 2-month protection that a single injection provides. For the main analysis we assumed the duration of this tail protection to be 6 months (LEN) and 3 months (CAB), based on available pharmacokinetic studies; we varied these in sensitivity analysis.^4,19^ The model assumed that the relative rates of uptake between the subpopulations modelled remains similar with injectable PrEP to that of oral TDF/FTC. More detail on the epidemiological model, including sources for assumptions, are available online at Thembisa.org.^13^

### Scenarios and assumptions

For the first analysis, we modelled LEN, CAB and TDF/FTC scale-up over a 20-year time horizon, starting from 2026, with uptake in all women, particularly adolescent girls and young women (AGYW) aged 15-24 years, pregnant and breastfeeding women (PBFW), FSW, MSM and heterosexual men. The baseline scenario was the current HIV programme with the current TDF/FTC roll-out, amounting to a coverage of 7% for AGYW, 0.1% (PBFW), 11% (FSW), 10% (MSM), and 0.2% (heterosexual men) (Table 1). For TDF/FTC and conservative LEN or CAB scale-up, we assumed initiation rates doubled compared to baseline, while under our optimistic LEN or CAB scale-up scenarios, we doubled these rates again. PBFW have low rates of PrEP uptake at baseline, but we assumed 29% coverage under TDF/FTC, 16% under LEN and 21%-22% under CAB. Under LEN and CAB scenarios we assumed continuation of current baseline rates of TDF/FTC initiation remain unchanged. We assumed an average duration of use for TDF/FTC of 3 months for all women and heterosexual men, and 6 months for MSM. Duration of use for LEN under the conservative scenario was assumed to be 6 months (women, heterosexual men) and 12 months (MSM), and under the optimistic scenario this doubled with 12 months (women, heterosexual men) and 24 months (MSM). Under CAB the duration assumed was less, with MSM having the longer assumed duration (conservative: 4-8 months; optimistic: 8-16 months). These assumptions resulted in varying coverage levels across scenarios and subpopulations, which are, along with other key assumptions, detailed in Table 1.

**Table 1.**
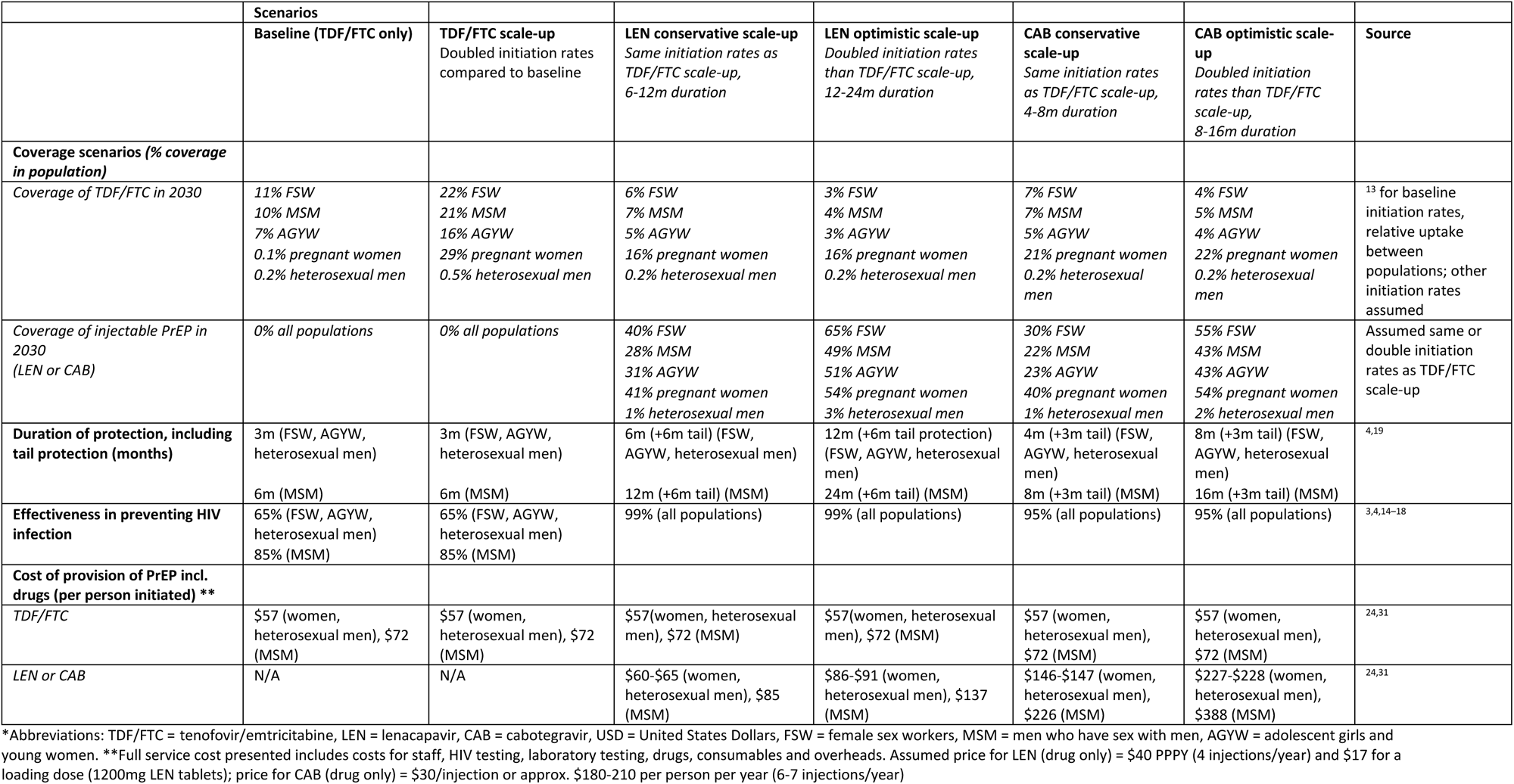
Key assumptions on coverage, duration and effectiveness for main scenarios.

For the second analysis, we compared the impact and cost per life-year saved of PrEP scale-up to expansion of other existing HIV interventions, similar to previous analysis for the annual South African HIV Investment Case.^20,21^ We constructed a simple league table by ranking interventions based on their incremental cost-effectiveness ratio (ICER) per life year saved. Interventions were scaled up separately, with each compared to baseline: ART (95% coverage, an increase from the current 81% coverage in 2025 among PLHIV who know their status), condom distribution (from 418 million to 725 million condoms/year), medical male circumcision (MMC) (from ~210,000/year to ~500,000/year), general HIV testing (from 16.5 million to 18 million), an HIV self-screening package consisting of distributing HIV self-test kits mostly to partners of ART patients (from 333,000/year to 1 million/year),^22^ PCR testing of infants at 10 weeks (from 80% to 95% coverage), PCR testing of infants at 6 months (from 40% to 95% coverage), and rapid HIV testing of infants at 18 months (from 30% to 95% coverage).

The third analysis modelled LEN strategies reaching different subpopulations to maximise the epidemiologic impact of (i) the current Global Fund LEN allocation (covering ~500,000 person-years over 2026-2027) (Phase 1),^6^ and (ii) the large-scale roll-out (Phase 2). The National Department of Health (NDOH) had indicated that the planned priority populations for Phase 1 were AGYW, PBFW, FSW, MSM and transgender individuals, reaching up to 500,000 initiations over 2026-2027. To model the impact of the Phase 1 allocation, we assumed no LEN initiations from 2028 onwards, and an average duration on LEN of one year for all populations, and simulated the new infections over five years 2026-2030, reflecting primary and short-term secondary infections averted in the total population. Transgender individuals were not included as Thembisa does not currently model this population dynamically. We modelled 490 combinations of discrete increments of LEN initiations across subpopulations, up to pre-defined maximum coverages of 60% for FSW (45,000 initiations/year) and 30% for MSM (93,500 initiations/year), and the total LEN available for AGYW and PBFW. (maximum 250,000 initiations/year, corresponding to ~4% and ~20% coverage, respectively) (Table S1). For Phase 2, we modelled a large-scale LEN roll-out delivered to each subpopulation separately, as well as selected combinations of AGYW, PBFW, FSW and MSM, over a 20-year time horizon (2026-2045), and compared the impact and cost to baseline. Our uptake assumptions were intentionally conservative, aligning with baseline TDF/FTC initiation numbers, and somewhat higher for key populations (FSW, MSM) to explore the potential impact under more optimistic uptake scenarios. We assumed conservative average durations: 6 months for women and heterosexual men, and 12 months for MSM. Overall this analysis resulted in a number of initiations for AGYW that could reach ~700,000/year, FSW: ~100,000/year, PBFW: ~400,000/year, MSM: ~150,000/year and heterosexual men: ~600,000/year.

### Cost analysis

Costs were analysed from the perspective of the South African government and reported in 2025 United States Dollar (USD; exchange rate 18.22 South African Rand (ZAR) per 1 USD).^23^ Costs are undiscounted, representing data of use for budgeting purposes. The cost of PrEP provision was estimated using an ingredients-based approach; the full methodology has been described elsewhere.^24^ Briefly, PrEP is provided in primary healthcare clinics and includes HIV testing, counselling, provision of condoms, syndromic management of sexually transmitted infections with treatment referral, training, outreach, mobilisation, monitoring and evaluation costs. Visit schedules varied according to each product (normalised to a 12-month duration; LEN: 2 per year; CAB: 7 per year; oral TDF/FTC: 5 per year). The cost of LEN was assumed to be $40 PPPY for the injections and $17 for the loading dose, as per the recently announced agreement with generic manufacturers and funders.^25^ The cost of CAB was $180 PPPY ($30 per injection),^26^ while oral TDF/FTC was $41 PPPY ($3.38 for one month’s supply) at current generic prices. The full-service cost of PrEP provision for all scenarios is summarised in Table 1. The estimation of costs of the other HIV interventions as well as the HIV programme overall followed the same approach as the South African HIV Investment Case.^20,27^ We estimated cost-effectiveness over a 20-year time horizon (2026-2045) as the incremental cost per HIV infection averted and incremental cost per life year saved, compared to baseline.

### Sensitivity analysis

We conducted a probabilistic sensitivity analysis to evaluate the impact of the uncertainty of 58 key parameters in Thembisa on the main cost-effectiveness analysis,^13^ including PrEP-specific parameters such as PrEP effectiveness, duration on TDF/FTC, tail protection for injectable PrEP, and reduction in condom use while on PrEP. We varied the cost of LEN service delivery (excluding drug costs) with lower and upper bounds varying by 50% from our average estimate. A Monte Carlo simulation, encompassing 1,000 iterations, sampled data from pre-determined distributions associated with PrEP-related key parameters as described in Table S2 in the supplementary material. We report median estimates across cost and health outcomes with 95% uncertainty bounds (UB) estimated using 2.5th and 97.5th percentiles.

### Ethics

As this study did not include primary human subjects data, no ethical clearance was sought.

## Results

### Epidemiological impact

The numbers initiating PrEP increased substantially in all modelled scenarios. Compared to the current number of TDF/FTC initiations of ~600,000/year, our TDF/FTC scale-up scenario and conservative CAB and LEN scenarios would increase initiations to up to 2.4 million initiations per year by 2045-an average of 1.7 million annually (Figure 1). The optimistic CAB and LEN scale-up scenarios required between 900,000 and 4.4 million initiations per year (average 2.9 million annually). As CAB and LEN have different dosing frequencies and assumptions regarding continuation duration, the number of doses required differs between scenarios (Figure 1).

**Figure 1.**
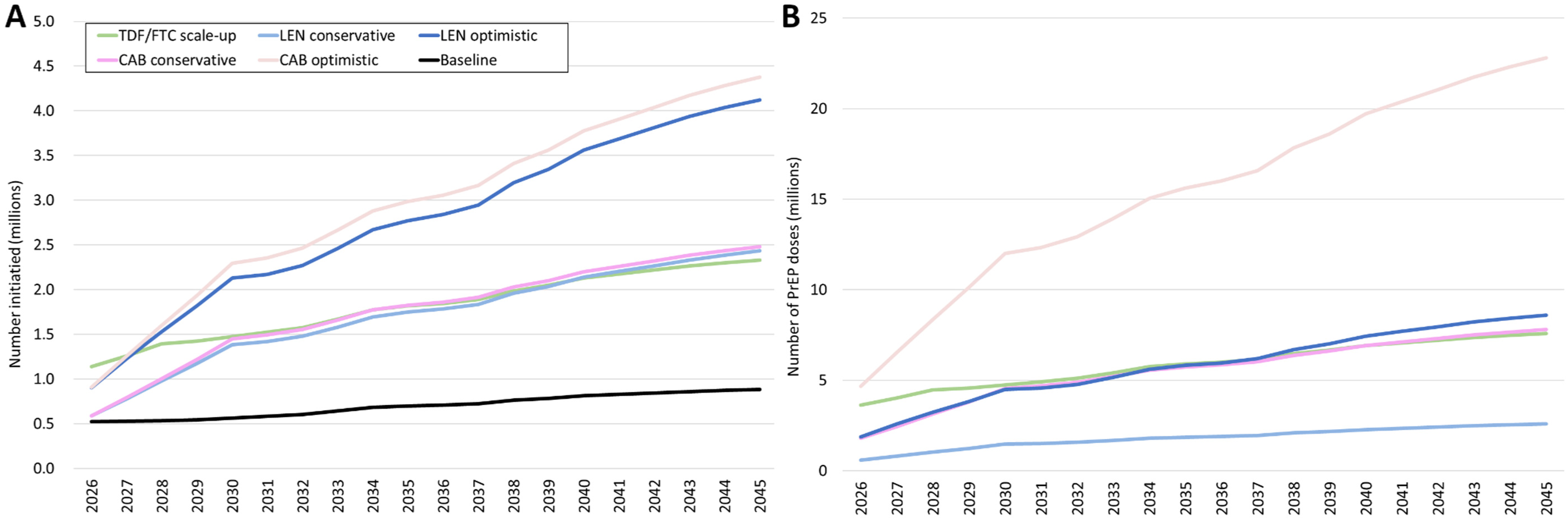
Number of (A) PrEP initiations and (B) doses by PrEP type per year for large-scale roll-out. Dosing frequency differs by PrEP type: TDF/FTC is represented by monthly packs; CAB requires an injection at initiation, month 1, and then 2-monthly; LEN requires an injection 6-monthly, loading dose for every initiation.

In the baseline scenario, population-wide HIV incidence was projected to continue declining and reach <0.1%, the virtual elimination threshold towards ending AIDS, by 2053. In the optimistic scenario, LEN scale-up accelerated this downward trajectory to reach <0.1% incidence by 2039, and avert on average 44,200 new HIV infections annually (Figure 2). Under conservative LEN scale-up, incidence reached <0.1% by 2043, and averted on average 26,900 infections/year (Figure 2). Injectable CAB averted slightly fewer infections, primarily because of the shorter continuation duration, with an average 35,500 (optimistic) and 19,500 (conservative) infections averted annually. CAB reached <0.1% incidence in 2042 (optimistic) and 2045 (conservative). In comparison, TDF/FTC scale-up averted an average of 6,300 infections/year, and only reached <0.1% incidence by 2050 (Figure 2). Compared to baseline, LEN averted 19%-31% of HIV infections, and CAB averted 14-25% of HIV infections over the 20-year period, and saved 593,000-989,300 (3-5%) life years, while CAB saved 428,700-792,600 (2-4%) life years (Table 2). The number of people needed to initiate PrEP to avert one HIV infection, after the initial 4-year scale-up period, was ~280 initiations for TDF/FTC, and considerably less under LEN (conservative: ~65; optimistic: ~35) and CAB (conservative: ~125; optimistic: ~45) (Figure S1).

**Figure 2.**
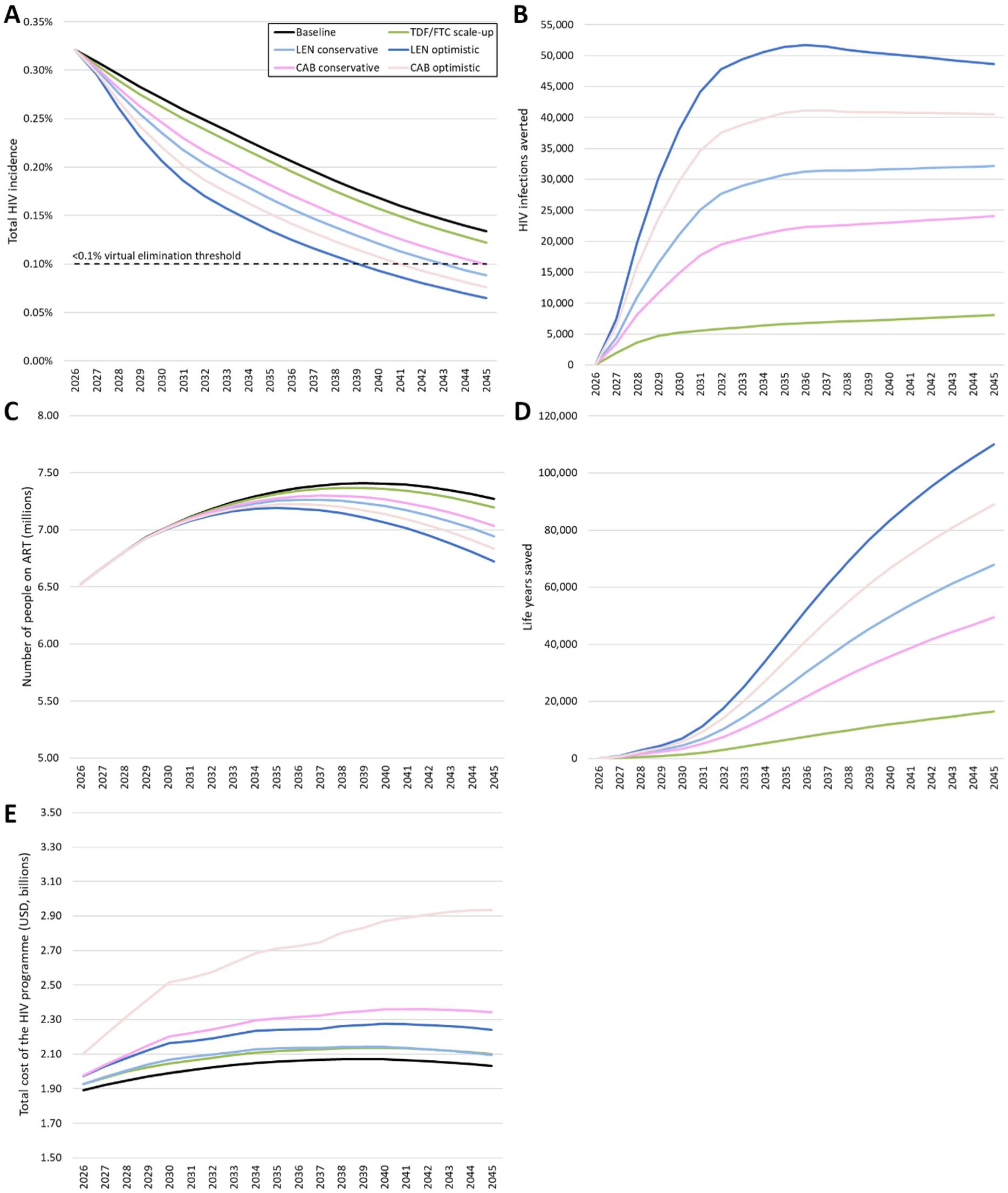
Impact of PrEP scale-up on (A) annual total HIV incidence, (B) HIV infections averted annually, (C) total number of people on ART, (D) life years saved over baseline, and (E) total HIV programme cost (in 2025 USD)

**Table 2.** Impact and cost-effectiveness of oral TDF/FTC, injectable LEN or CAB scale-up compared with baseline over 2026–45.

| Scenario | Total Cost of the HIV programme |  | Incremental cost effectiveness |  | New HIV infections |  | Life years lost due to AIDS |  |
| --- | --- | --- | --- | --- | --- | --- | --- | --- |
|  | Cost (billions, USD) | Incremental cost over baseline, % | Cost per infection averted (USD) | Cost per life year saved (USD) | Number (millions) | % averted over baseline | Number (millions) | % saved over baseline |
| **Baseline** | 40.49 | - | - | - | 2.72 | - | 19.89 | - |
| **Oral TDF/FTC scale up** | 41.68 | 3% | \$9,907 | \$8,143 | 2.60 | 4% | 19.74 | 1% |
| **Lenacapavir** |  |  |  |  |  |  |  |  |
| Conservative | 41.85 | 3% | \$2,666 | \$2,301 | 2.21 | 19% | 19.29 | 3% |
| Optimistic | 44.02 | 9% | \$4,202 | \$3,567 | 1.88 | 31% | 18.90 | 5% |
| **Cabotegravir** |  |  |  |  |  |  |  |  |
| Conservative | 45.25 | 12% | \$12,877 | \$11,114 | 2.35 | 14% | 19.46 | 2% |
| Optimistic | 53.26 | 32% | \$18,918 | \$16,118 | 2.05 | 25% | 19.09 | 4% |

Abbreviations: TDF/FTC = tenofovir/emtricitabine, LEN = lenacapavir, CAB = cabotegravir, USD = United States Dollars, ART=antiretroviral treatment, PPPY=per person per year

### Cost and cost effectiveness of LEN and CAB

The cost of TDF/FTC provision was $57-$72 per person initiated, while the cost of LEN and CAB provision ranged from $60-$137 and $146-$388 per person initiated, respectively. Costs varied by population and were highly dependent on the assumed duration on product. The primary cost driver was drugs for LEN (65%) and CAB (69%), while staff costs (36%) were the largest component for TDF/FTC (Table S3). Both TDF/FTC and conservative LEN scale-up increased total HIV programme cost by 3%; however, LEN resulted in substantially greater health impact, rendering it more cost-effective (Table 2). The ICER of scaling-up LEN was $2,666-$4,202/infection averted and $2,301-$3,567/life year saved (LYS), compared to TDF/FTC with $9,907/infection averted and $8,143/LYS (Table 2). CAB was less cost-effective than both LEN and oral TDF/FTC scale-up with an ICER of $12,877-$18,918/infection averted and $11,114-$16,118/LYS over baseline (Table 2).

Compared to further scaling-up other HIV interventions, LEN had a larger impact on both life years saved and infections averted than most, second only to scaling up ART coverage to 95% (Table 3). However, it was only moderately cost-effective, ranking 6^th^ (conservative) and 8^th^ (optimistic) among modelled interventions when comparing ICER/LYS (Table 3). Interventions more cost-effective than LEN under this metric were scaling-up condoms (cost-saving), ART coverage, HIV testing of the general population, MMC and HIV self-screening, with ICERs ranging from $577-$1,081/LYS. In terms of ICER/infection averted, LEN ranked fourth, but remained less cost-effective than condoms, MMC, and ART (Table 3).

**Table 3.** Simple league table comparing 20-year impact (2026-2045) and cost-effectiveness of LEN and CAB scale-up scenarios to existing HIV interventions, with incremental cost effectiveness ratios compared to baseline.

| Intervention | Incremental cost over baseline, USD millions (% change) | Incremental life years saved over baseline (% change) | Incremental HIV infections averted over baseline (% change) | ICER per life year saved (USD) | Rank (ICER/ LYS) | ICER per HIV averted (USD) | Rank (ICER/ HIV) |
| --- | --- | --- | --- | --- | --- | --- | --- |
| Condom distribution (725m/year) | -301 (-0.7%) | 348,165 (1.8%) | 260,263 (9.6%) | Cost-saving | 1 | Cost-saving | 1 |
| ART (95%) | 2,752 (6.8%) | 4,770,659 (24.0%) | 1,414,990 (52.0%) | 577 | 2 | 1,945 | 3 |
| General HTS (18m/year) | 149 (0.4%) | 167,249 (0.8%) | 38,844 (1.4%) | 889 | 3 | 3,827 | 5 |
| MMC (500k/year) | 103 (0.3%) | 101,903 (0.5%) | 101,324 (3.7%) | 1,007 | 4 | 1,012 | 2 |
| HIV self-screening package (1m/year) | 213 (0.5%) | 196,712 (1.0%) | 35,935 (1.3%) | 1,081 | 5 | 5,917 | 7 |
| LEN conservative (avg 1.8m/year) | 1,365 (3.4%) | 593,025 (3.0%) | 511,955 (18.8%) | 2,301 | 6 | 2,666 | 4 |
| Infant PCR testing at 10 weeks (95%) | 18 (<0.1%) | 6,412 (<0.1%) | 17 (<0.1%) | 2,850 | 7 | 1,074,866 | 12 |
| LEN optimistic (avg 2.9m/year) | 3,529 (8.7%) | 989,334 (5.0%) | 839,898 (30.9%) | 3,567 | 8 | 4,202 | 6 |
| Oral TDF/FTC (avg 1.8m/year) | 1,189 (2.9%) | 145,977 (0.7%) | 119,982 (4.4%) | 8,143 | 9 | 9,907 | 8 |
| Infant rapid HIV testing at 18 months (95%) | 76 (0.2%) | 8,770 (<0.1%) | 496 (<0.1%) | 8,645 | 10 | 152,862 | 11 |
| CAB conservative (avg 1.8m/year) | 4,764 (11.8%) | 428,674 (2.2%) | 369,997 (13.6%) | 11,114 | 11 | 12,877 | 9 |
| CAB optimistic (avg 2.9m/year) | 12,775 (31.6%) | 792,593 (4.0%) | 675,315 (24.8%) | 16,118 | 12 | 18,918 | 10 |
| Infant PCR testing at 6 months (95%) | 66 (0.2%) | 1,484 (<0.1%) | 55 (<0.1%) | 44,192 | 13 | 1,192,390 | 13 |

### Budget impact

The conservative LEN scale-up scenario required an additional $38-$91 million/year (2-5% over the current projected HIV programme cost) over financial years 2025/26-2029/30 (Table S4). This incremental cost was similar to the TDF/FTC scale-up scenario, but with a substantially larger impact (Table 2). Under an optimistic scenario, $84-$198 million/year (4%-10% increase) was required for scale-up (Table S4).

### Optimization of LEN to subpopulations and most cost-effective long-term strategies

Optimised allocation of 500,000 PYs of LEN delivered in 2026-2027 could avert a maximum of 20,600 infections over 2026-2030. The optimised allocation meant that 55% of LEN doses would be allocated to PBFW, 26% to MSM and 18% to FSW (Figure 3); this averted ~20,500 new infections over 2026-2030, relative to baseline. However, alternative allocation strategies that included AGYW had comparable impact. When incrementally shifting LEN initiation towards AGYW (10% increments representing ~25,000 initiations annually), LEN would avert between 18,100 and 20,000 infections with 10-50% of doses allocated to AGYW (Table S5). Increasing LEN initiations in AGYW maintains strong impact, but reduces PBFW coverage and overall impact. For a large-scale roll-out, the most cost-effective strategies were delivering to FSW only, MSM only, or FSW and MSM concurrently, with ICERs $276-$958/LYS over a 20-year time horizon (Table 4). Amongst strategies modelled, the combined FSW and MSM approach averted the most infections (~187,000) over the 20 years. Delivering to a combination of subpopulations that include PBFW and AGYW resulted in higher ICER values ranging $1,769-$3,020/LYS, while LEN to heterosexual men was the least cost-effective strategy ($9,806/LYS) (Table 4).

**Figure 3.**
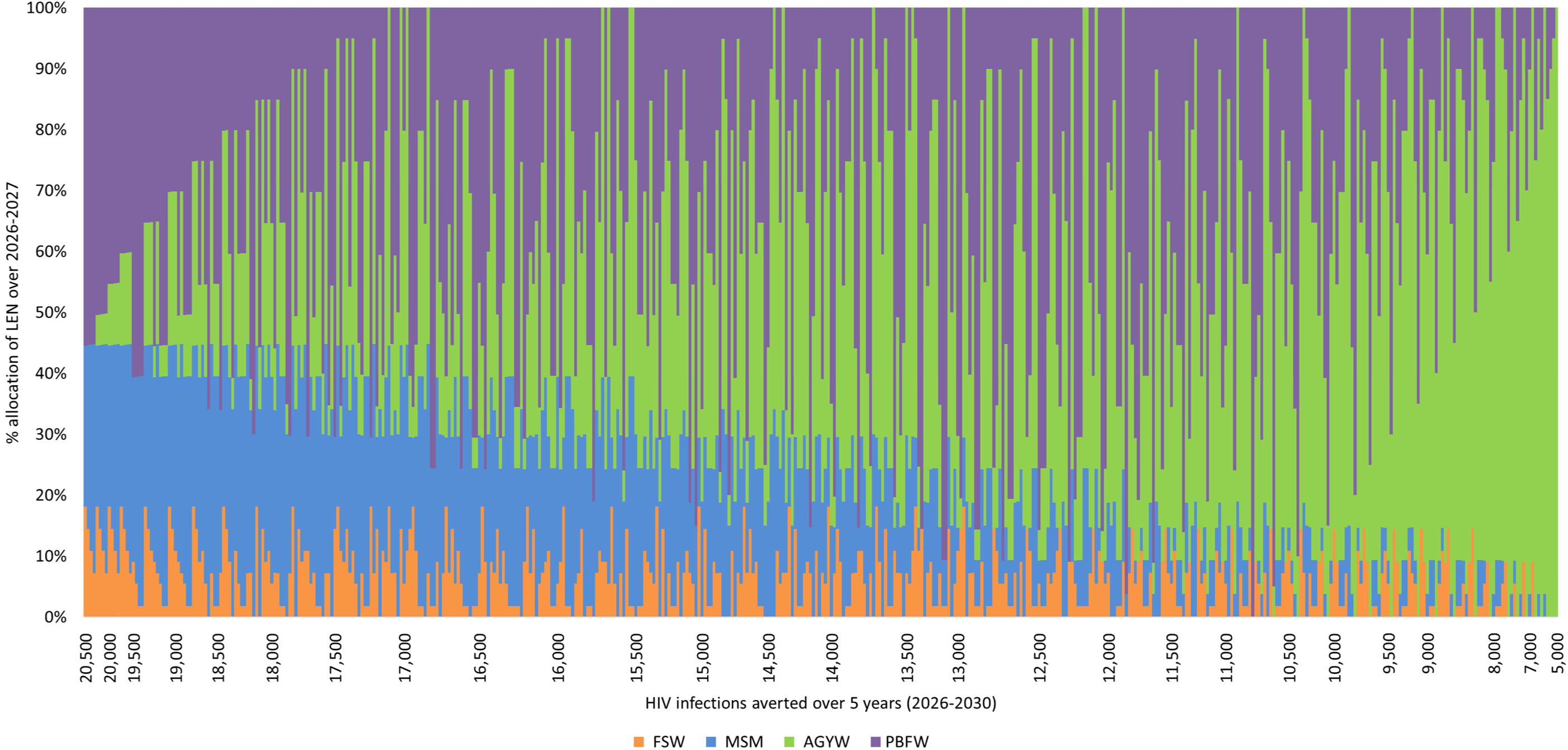
Five-year impact of different combinations of subpopulation uptake of LEN for initial allocation volume (~500,000 person-years on LEN) over 2026-2027. Each of the 490 different combinations of LEN distribution are represented by vertical bars, with all results sorted by descending order of impact, therefore the leftmost combinations represent the highest impact strategies.

**Table 4.** Impact and cost-effectiveness of specific sub-population uptake of LEN, compared to baseline, over 20 years (2026-2045)

| **Scenario** | **Total cost of the HIV programme**<br>**(USD, billions)** | **Incremental cost over baseline**<br>**(USD, millions)** | **Life years lost to AIDS**<br>**(millions)** | **Total new HIV infections**<br>**(millions)** | **Incremental life years saved**<br>**(thousands)** | **Incremental HIV infections averted**<br>**(thousands)** | **ICER per life year saved**<br>**(USD)** | **ICER per HIV averted**<br>**(USD)** |
| --- | --- | --- | --- | --- | --- | --- | --- | --- |
| Baseline | 40.49 | - | 19.89 | 2.72 | - | - | - | - |
| FSW | 40.51 | 17 (0.04%) | 19.82 | 2.67 | 62 (0.3%) | 52 (1.9%) | 276 | 332 |
| FSW and MSM | 40.66 | 174 (0.4%) | 19.67 | 2.53 | 216 (1.1%) | 187 (6.9%) | 804 | 928 |
| MSM | 40.64 | 155 (0.4%) | 19.72 | 2.58 | 162 (0.8%) | 143 (5.2%) | 958 | 1,086 |
| FSW, MSM and pregnant women | 41.12 | 633 (1.6%) | 19.53 | 2.44 | 358 (1.8%) | 286 (10.5%) | 1,769 | 2,216 |
| FSW and AGYW | 41.05 | 560 (1.4%) | 19.60 | 2.45 | 282 (1.4%) | 272 (10.0%) | 1,982 | 2,056 |
| AGYW | 41.03 | 540 (1.3%) | 19.65 | 2.49 | 232 (1.2%) | 231 (8.5%) | 2,332 | 2,339 |
| Pregnant women | 40.94 | 454 (1.1%) | 19.74 | 2.62 | 150 (0.8%) | 106 (3.9%) | 3,020 | 4,284 |
| Heterosexual men | 41.21 | 719 (1.8%) | 19.81 | 2.67 | 73 (0.4%) | 50 (1.8%) | 9,806 | 14,458 |

### Sensitivity analysis

The relative impact of PrEP scenarios was not sensitive to uncertainty in epidemic calibration and PrEP intervention parameters; the reduction of HIV infections for LEN (conservative UB: 13.8%-24.1%; optimistic UB: 25.5%-35.7%) and CAB (conservative UB: 10.0%-18.8%; optimistic UB: 20.1%-30.0%) still exceeded TDF/FTC (UB 3.4%-5.5%) (Table S6). LEN remained the most cost-effective option with consistently lower ICERs but wider uncertainty (conservative UB: $803-$7,142/LYS; optimistic UB: $1,761-$9,897), compared to TDF/FTC (UB $5,863-$19,157/LYS) and CAB (conservative UB: $6,516-$28,606/LYS; optimistic UB: $10,455-$38,496) (Figure S2). CAB was consistently less cost-effective than LEN in all simulations, and less cost-effective than TDF/FTC scale-up in 88% (conservative) and 99.8% (optimistic) of simulations (Figure S2).

## Discussion

Widespread use of LEN among those with elevated exposure to HIV could substantially decrease HIV infections and accelerate efforts to end AIDS in South Africa, achieving the <0.1% incidence threshold required to end AIDS 10-14 years earlier than through continuing current intervention levels, and 7-11 years sooner than further aggressive TDF/FTC scale-up. At $40 PPPY and $17 for the loading dose and an average delivery cost of $30 PPPY, LEN scale-up options were more cost-effective than both TDF/FTC and CAB scale-up but increased the HIV programme medium-term budget by 4-8%. CAB was the least cost-effective option, due to its high price.

While the newly negotiated price for LEN substantially improves affordability, large-scale roll-out would still require substantial additional investment, which may particularly strain national funding in the current context of reduced US foreign aid. Compared to other interventions, expanding ART to 95% coverage of all diagnosed PLHIV was the only intervention exhibiting greater cost-effectiveness than LEN and higher impact in terms of both life years saved and HIV infections averted. However, significant implementation challenges remain in achieving 95% ART coverage, and the feasibility and resource requirements of reaching this target are not well established. Although we did not explicitly compare interventions to budget constraints here, previous results imply that if HIV programme resources are threatened, maintaining people on ART should be safeguarded before scaling-up LEN.^28^ Increasing condom distribution and MMC also remained more cost-effective prevention options than LEN and should be implemented alongside PrEP scale-up as part of a comprehensive HIV prevention strategy for South Africa; however, the overall incremental epidemiologic impact of further expanding these interventions was limited compared to LEN.

Similar to other modelling analyses, we assumed PrEP uptake would be risk differentiated, reaching individuals at disproportionately higher risk among AGYW, PBFW, and heterosexual men. Even with universal access, such differentiated uptake has defined efficient and impactful oral PrEP programmes in some settings,^12^ but evidence is less clear in African epidemic settings. Some of our findings may be sensitive to this assumption, including the projected magnitude of infections averted, cost-effectiveness, and recommending prioritisation of PBFW versus AGYW. This prioritisation could change if, for example, delivery models reaching wider AGYW (e.g. through STI and other sexual and reproductive health services) were more efficient at reaching those at highest risk than delivery to pregnant women at ANC services, where providers may find it more challenging to differentiate clients.

Ambitious LEN coverage modelled among FSW and MSM may be difficult to attain, particularly in the current context. Termination of US donor funding has disproportionately affected key population services in South Africa, which were primarily managed by organizations supported by PEPFAR or the Global Fund. Key populations often face stigma, which complicates their utilization of public health facilities, while PBFW are already served within public clinics that can provide LEN. Successful delivery of LEN to FSW and MSM will likely require the South African government to revive key population programming in a way that provides friendly and safe care, potentially through partnering with existing experienced implementing organizations. In addition, geospatial prioritisation of areas with the highest HIV incidence could further enhance both the cost-effectiveness and overall impact of LEN implementation at scale.^29^

Our costing approach differs from previous modelling studies of LEN in the region, with Wu *et al.* and Kaftan *et al.* using a provision cost of $8.55 per dose from a study in Kenya.^30^ This cost is based on a streamlined 6-month dispensing model of oral PrEP with interim HIV self-testing. While this approach is valid for modelling efficiency gains, it may underestimate the costs of implementing new injectable products at scale in routine settings. Our ingredients-based approach resulted in an average provision cost of $30 PPPY (excluding drugs) in the routine public-sector clinics, and indicates that, despite these higher delivery cost estimates, LEN remains highly cost-effective at the negotiated price of $40 PPPY, suggesting its value proposition is robust under conservative costing assumptions.

Our analysis is subject to several limitations. First, the feasibility of delivering our modelled scenarios is currently unknown. We included costs for demand creation, but these may not be sufficient to reach the assumed large-scale uptake. In addition, South Africa may need to re-establish key population services in order to target those populations, and this may require additional resources. Together, these factors suggest that we may be overestimating LEN’s cost-effectiveness. However, our conservative scenario assumes most clients receive only one injection (two for MSM), which may partially mitigate this concern. Second, with limited data available on costs of implementing of long-acting products, we opted for an ingredients-based approach. The biggest cost difference between PrEP options lies in the drugs, but further research is needed to determine the true implementation costs and explore innovative delivery methods for injectable PrEP to enhance its cost-effectiveness, especially amongst key populations. Third, our modelling assumed continuation of a baseline trajectory and intervention coverages that pre-date recent funding cuts. While the relative comparisons between interventions and overall conclusions remain valid, key populations were disproportionately affected by funding cuts and may have experienced increased HIV incidence as a result of disrupted prevention services.^28^ Higher incidence would translate to greater reductions of HIV if effective prevention were scaled up in these populations.

Immediate and concurrent monitoring and evaluation of the upcoming LEN roll-out are essential to generate evidence of uptake patterns and preferences, especially across different subpopulations. Such evidence is needed to inform efficient scale-up strategies as the PrEP product landscape continues to expand. As the LEN roll-out and continued insights will help guide national decision-making, South Africa’s scale and epidemic burden mean it is not merely a participant in the PrEP market – it is a market shaper whose adoption choices will influence global pricing and supply trajectories.

## Data sharing

The C++ version of Thembisa is available on a GitHub repository at https://github.com/leighjohnson/Thembisa

## Contributors

LJ, LFJ, JWI-E and GMR conceptualised the analytical framework and scenarios. LJ and GMR developed the unit costs and drafted the manuscript. LFJ developed the epidemiological model. LJ performed the modelling and economic analyses. LJ and LFJ accessed and verified all data. LGB provided trial data and key information regarding implementation to inform cost and epidemiological assumptions. HS consulted on the NDOH plan for roll-out and provided valuable context to inform our approach. All authors critically reviewed and approved the final manuscript, and share final responsibility for the decision to submit for publication. All authors had access to all data in the study.

