## Supplementary for "Taking injectable PrEP to scale: Optimising the value of lenacapavir for South Africa’s HIV response"

**Table S1. Subpopulation prioritization: number of initiations assumed for each population.**

All possible combinations of the below targets modelled where the total number of initiations ranged between 246,000 and 250,000 per year (totalling to 492,000-500,000 initiations over 2026/27). All populations are assumed to remain on LEN for 12 months. FSW and MSM were capped at a maximum coverage of 60% and 30% of HIV negative populations, respectively.

| **Adolescent girls and young women (AGYW)** | **Pregnant/breastfeeding women (PBFW)** | **Female sex workers (FSW)** | **Men who have sex with men (MSM)** |
| --- | --- | --- | --- |
| 0 | 0 | 0 | 0 |
| 12,500 | 12,500 | 4,500 | 9,360 |
| 25,000 | 25,000 | 9,000 | 18,720 |
| 37,500 | 37,500 | 13,500 | 28,080 |
| 50,000 | 50,000 | 18,000 | 37,440 |
| 62,500 | 62,500 | 22,500 | 46,800 |
| 75,000 | 75,000 | 27,000 | 56,160 |
| 87,500 | 87,500 | 36,000 | 65,520 |
| 100,000 | 100,000 | 45,000 | 74,880 |
| 112,500 | 112,500 |  | 84,240 |
| 125,000 | 125,000 |  | 93,600 |
| 137,500 | 137,500 |  |  |
| 150,000 | 150,000 |  |  |
| 162,500 | 162,500 |  |  |
| 175,000 | 175,000 |  |  |
| 187,500 | 187,500 |  |  |
| 200,000 | 200,000 |  |  |
| 212,500 | 212,500 |  |  |
| 225,000 | 225,000 |  |  |
| 237,500 | 237,500 |  |  |
| 250,000 | 250,000 |  |  |

**Table S2. Probability distributions used for parameters varied in the probabilistic sensitivity analysis**

| **Variable** | **Population** | **Distribution** | **Mean,**  **standard**  **deviation** |
| --- | --- | --- | --- |
| Reduction in condom use while on PrEP | All populations | Beta (0.80, 7.20) | 0.10, 0.10 |
| TDF/FTC effectiveness | Women and heterosexual men | Beta (14.14,7.61) | 0.65, 0.10 |
|  | Pregnant women | Beta (10.69,13.06) | 0.65, 0.10 |
|  | MSM | Beta (9.99, 1.76) | 0.85, 0.10 |
| CAB effectiveness | All populations | Beta (111.86, 5.89) | 0.95, 0.05 |
| CAB tail protection (months) | All populations | Gamma (2.25,0.75) | 3.0, 2.0 |
| LEN effectiveness | All populations | Beta (69.11, 1.12) | 0.984, 0.014 |
| LEN tail protection (months) | All populations | Gamma (2.25,0.38) | 6.0, 4.0 |
| Cost of LEN 6-12-month duration (including drugs) | Women (6 months) | Uniform ($42, $87) | $65, $13 |
|  | Heterosexual men (6 months) | Uniform ($40, $80) | $60, $12 |
|  | MSM (12 months) | Uniform ($63, $108) | $85, $13 |
| Cost of LEN 12-24-month duration (including drugs) | Women (12 months) | Uniform ($66, $117) | $91, $15 |
|  | Heterosexual men (12 months) | Uniform ($63, $109) | $86, $13 |
|  | MSM (24 months) | Uniform ($109, $166) | $137, $17 |
| Cost of CAB 4-8-month duration (including drugs) | Women (4 months) | Uniform ($124, $170) | $147, $13 |
|  | Heterosexual men (4 months) | Uniform ($123, $168) | $146, $13 |
|  | MSM (8 months) | Uniform ($197, $255) | $226, $17 |
| Cost of CAB 8-16-month duration (including drugs) | Women (8 months) | Uniform ($198, $259) | $228, $18 |
|  | Heterosexual men (8 months) | Uniform ($197, $257) | $227, $17 |
|  | MSM (16 months) | Uniform ($345, $432) | $388, $25 |

We ran 1,000 Monte Carlo simulations, sampling the variables from their distributions and shape parameters in each run. Table S2 summarizes the distributions from which the PrEP parameters are sampled in the uncertainty analysis. In addition to these, we sample from the posterior distributions generated in the calibration of the Thembisa model to various HIV data sources, so that the uncertainty ranges also reflect uncertainty regarding epidemiological processes. These include the uncertainty in HIV incidence trends, the relative HIV incidence across age/sex/risk groups, rates of HIV disease progression and mortality, and rates of HIV testing and ART linkage. More details regarding the specific parameters and their posterior distributions are included in the Thembisa 4.8 report^[1]^: Table 8.1 of the report summarizes the posterior distributions for adult HIV transmission and disease progression, Table B9 summarizes the posterior distributions for the HIV testing parameters, Table E6 summarizes the posterior distributions for the paediatric HIV parameters, and section 7.4 summarizes the distributions used to represent uncertainty in the future ART coverage, medical male circumcision uptake and condom use.

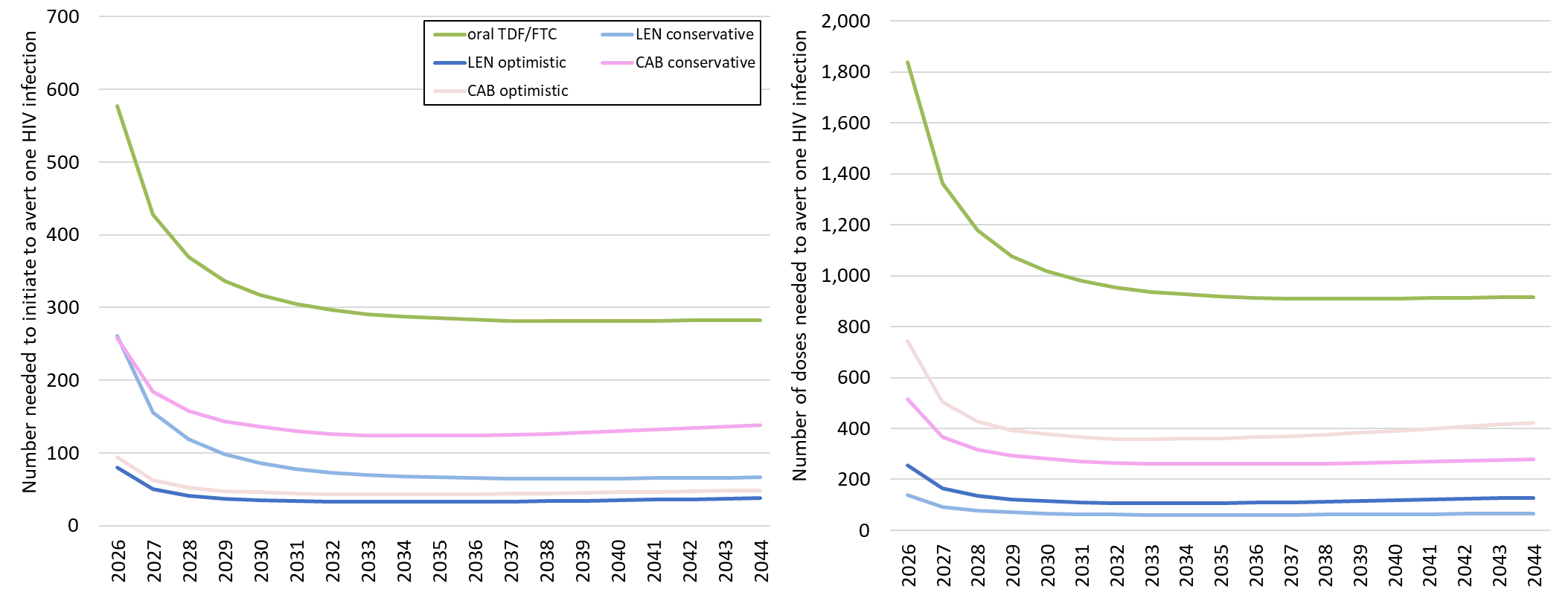
**Figure S1. (A) Number of people needed to initiate, and (B) doses required, to avert one HIV infection**

**B**

**A**

**Figure S2. Probabilistic sensitivity analysis results comparing the incremental cost per life year saved over 2026-2045 across 1,000 simulations for TDF/FTC scale-up to that of the LEN (A) conservative and (B) optimistic scale-up scenarios, and the CAB (C) conservative, and (D) optimistic scale-up scenarios**

*Each dot represents a single simulation, the diagonal line represents equal cost-effectiveness between TDF/FTC and LEN/CAB. The price of LEN was $40 PPPY (4 injections) and $17 for the loading dose tablets; price of CAB is the current manufacturer offered price of $180-210 PPPY (6-7 injections)*

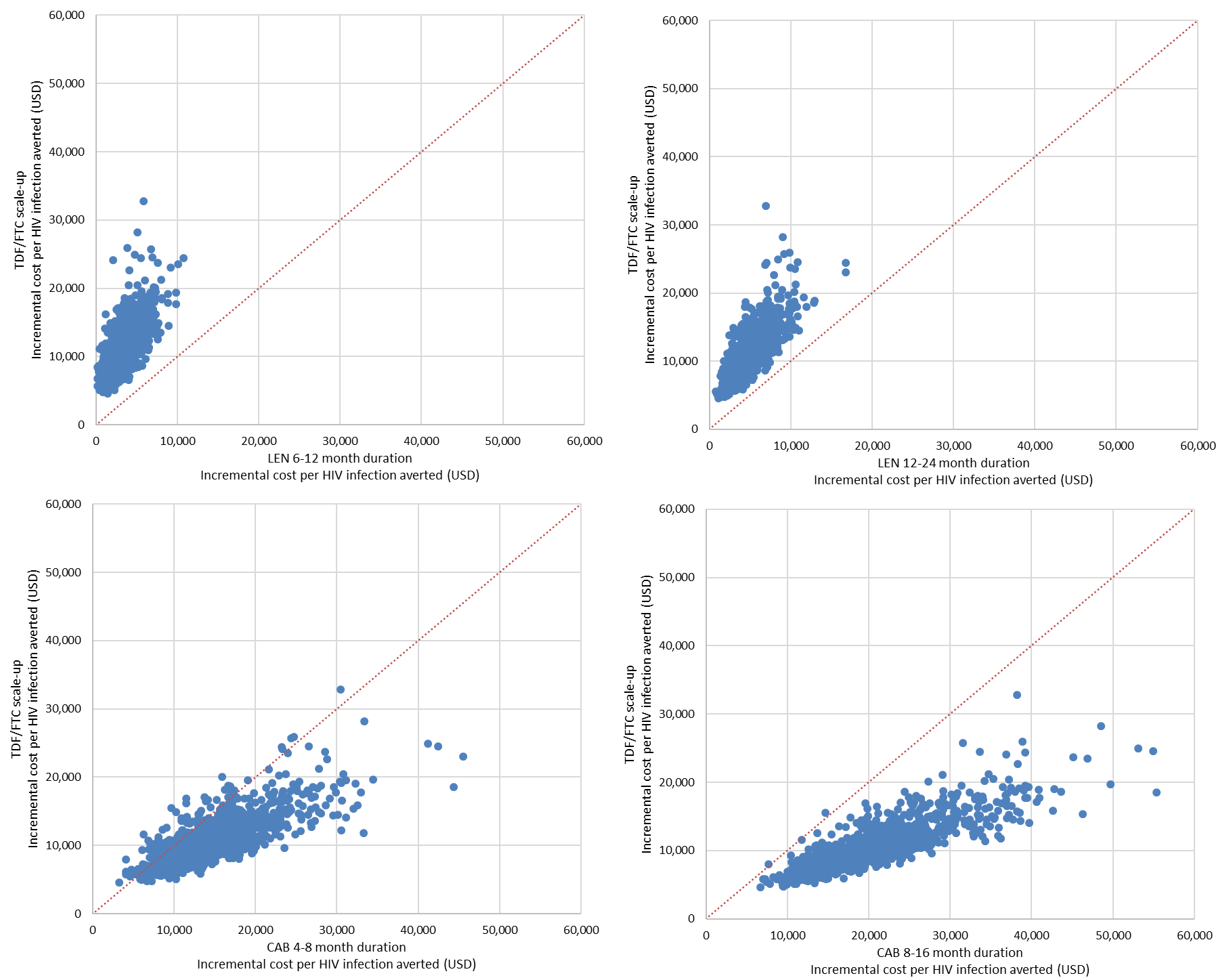

**B**

**A**

**D**

**C**

**Table S3. Cost breakdown of each PrEP modality by population and duration**

| **Cost of LEN provision** | | | | | | |
| --- | --- | --- | --- | --- | --- | --- |
| **Cost category** | **Women** | | **Heterosexual men** | | **MSM** | |
|  | **6 months duration** | **12 months duration** | **6 months duration** | **12 months duration** | **12 months duration** | **24 months duration** |
| Drugs | $37.00 | $57.00 | $37.00 | $57.00 | $57.00 | $97.00 |
| Staff | $20.02 | $23.63 | $15.80 | $19.40 | $19.63 | $26.87 |
| HIV tests | $3.11 | $3.81 | $3.14 | $3.83 | $2.41 | $3.81 |
| Consumables | $0.47 | $0.85 | $0.20 | $0.30 | $0.33 | $0.53 |
| Overheads | $4.39 | $6.12 | $4.08 | $5.78 | $5.70 | $9.12 |
| **Total** | **$64.99** | **$91.40** | **$60.22** | **$86.32** | **$85.07** | **$137.33** |
| **Cost of TDF/FTC provision** | | | | | | |
| **Cost category** | **Women (3 months duration)** | | **Heterosexual men (3 months duration)** | | **MSM (6 months duration)** | |
| Drugs | $13.52 | | $13.52 | | $23.66 | |
| Staff | $20.70 | | $20.02 | | $23.73 | |
| Laboratory testing | $13.98 | | $14.75 | | $14.57 | |
| Consumables | $0.96 | | $0.55 | | $0.96 | |
| Overheads | $8.50 | | $8.47 | | $9.46 | |
| **Total** | **$57.65** | | **$57.32** | | **$72.38** | |
| **Cost of CAB provision** | | | | | | |
| **Cost category** | **Women** | | **Heterosexual men** | | **MSM** | |
|  | **4 months duration** | **8 months duration** | **4 months duration** | **8 months duration** | **8 months duration** | **16 months duration** |
| Drugs | $100.54 | $167.56 | $100.54 | $167.56 | $167.56 | $301.61 |
| Staff | $27.24 | $34.47 | $27.25 | $34.46 | $34.73 | $49.21 |
| Laboratory testing | $7.80 | $9.19 | $7.82 | $9.20 | $7.80 | $10.60 |
| Consumables | $1.30 | $2.11 | $0.48 | $0.73 | $0.76 | $1.27 |
| Overheads | $9.73 | $15.08 | $9.67 | $14.98 | $14.91 | $25.53 |
| **Total** | **$146.61** | **$228.41** | **$145.76** | **$226.93** | **$225.77** | **$388.23** |

**Table S4. Budget impact analysis of LEN scale-up**

*All costs include service provision and drugs, and are presented in 2025 USD.*

| **Conservative lenacapavir scale-up, same initiation rates as TDF/FTC, 6-12-month duration** | | | | | |
| --- | --- | --- | --- | --- | --- |
|  | **2025/26** | **2026/27** | **2027/28** | **2028/29** | **2029/30** |
| Clients initiated (millions) | 0.59 | 0.78 | 0.98 | 1.17 | 1.39 |
| Doses required (millions) | 0.61 | 0.82 | 1.03 | 1.24 | 1.47 |
| **Total cost in USD, millions (% increase over current HIV programme cost)** | 38 (2%) | 51 (3%) | 64 (3%) | 77 (4%) | 91 (5%) |
| *Drugs: $40 for injections PPPY (2 x 927mg); $17 per 4x300mg loading dose tablets*  *Cost of provision (incl drugs) per person initiated: $85 (MSM); $60-$65 (women, heterosexual men)* | | | | | |
| **Optimistic lenacapavir scale-up, higher initiation rates than TDF/FTC, 12-24-month duration** | | | | | |
|  | **2025/26** | **2026/27** | **2027/28** | **2028/29** | **2029/30** |
| Clients initiated (millions) | 0.91 | 1.23 | 1.53 | 1.82 | 2.13 |
| Doses required (millions) | 1.88 | 2.59 | 3.23 | 3.84 | 4.50 |
| **Total cost in USD, millions (% increase over current HIV programme cost)** | 84 (4%) | 115 (6%) | 143 (7%) | 169 (9%) | 198 (10%) |
| *Drugs: $40 for injections PPPY (2 x 927mg); $17 per 4x300mg loading dose tablets*  *Cost of provision (incl drugs) per person initiated: $137 (MSM); $86-$91 (women, heterosexual men)* | | | | | |

**Table S5. Selected distribution scenarios of LEN allocation of ~500,000 person-years on LEN over 2026-27 for maximum impact on HIV infections**

|  |  | **% of LEN allocation distributed to subpopulations**  *(number of person-years on LEN)* | | | |
| --- | --- | --- | --- | --- | --- |
| **Scenario** | **Infections averted**  **over 2026-2030** | **FSW** | **MSM** | **AGYW** | **PBFW** |
| *Overall highest impact* | **20,500** | 18.1% *(90,718)* | 26.4% (*132,086*) | 0.0% (*0*) | 55.4% (*277,195*) |
| **Highest impact at set distribution to AGYW** | | | | | |
| *~10% to AGYW* | **20,000** | 18.1% *(90,718)* | 26.4% (*132,086*) | 10.1% (*50,399*) | 45.4% (*226,796*) |
| *~20% to AGYW* | **19,400** |  |  | 20.2% (*100,798*) | 35.3% (*176,397*) |
| *~30% to AGYW* | **18,800** |  |  | 30.2% (*151,197*) | 25.2% (*125,998*) |
| *~40% to AGYW* | **18,100** |  |  | 40.3% (*201,597*) | 15.1% (*75,599*) |
| *~50% to AGYW* | **17,400** |  |  | 50.4% (*251,996*) | 5.0% (*25,200*) |

**Table S6. Median, lower and upper uncertainty bounds around the impact and cost-effectiveness of TDF/FTC, LEN and CAB over a 20-year time horizon (2026-2045); based on 1,000 Monte Carlo simulations in a probabilistic sensitivity analysis*;** *figures represent median estimate across simulations and in brackets 2.5^th^ and 97.5^th^ percentiles.*

|  | **Total Cost of the HIV programme** | | **Incremental cost effectiveness** | | | **New HIV infections** | | | **Life years lost due to AIDS** | | |
| --- | --- | --- | --- | --- | --- | --- | --- | --- | --- | --- | --- |
| **Scenario** | **Cost**  **(billions, USD)** | **Incremental cost over baseline, %** | | **Cost per infection averted (USD)** | **Cost per life year saved (USD)** | | **Number (millions)** | **% averted over baseline** | | **Number (millions)** | **% saved over baseline** |
| **Baseline** | 39.77 (37.02-43.27) | - | | - | - | | 2.61 (1.78-3.83) | - | | 19.28 (16.52-22.96) | - |
| **Oral TDF/FTC scale up** | 40.97 (38.42-44.29) | 3.0% (2.4%-3.8%) | | 10,336 (5,863-19,157) | 8,445 (4,958-15,023) | | 2.50 (1.71-3.67) | 4.4% (3.4%-5.5%) | | 19.14 (16.42-22.78) | 0.7% (0.5%-0.9%) |
| **Lenacapavir** |  |  | |  |  | |  |  | |  |  |
| Conservative | 41.22 (38.60-44.40) | 3.6% (2.6%-4.3%) | | 2,930 (803-7,142) | 2,539 (669-5,904) | | 2.12 (1.48-3.08) | 18.1% (13.8%-24.1%) | | 18.70 (16.18-22.18) | 2.8% (1.9%-4.1%) |
| Optimistic | 43.45 (40.86-46.29) | 9.2% (7.0%-10.4%) | | 4,481 (1,761-9,897) | 3,803 (1,530-8,072) | | 1.81 (1.29-2.58) | 30.4% (25.5%-35.7%) | | 18.32 (15.91-21.61) | 4.9% (3.5%-6.3%) |
| **Cabotegravir** |  |  | |  |  | |  |  | |  |  |
| Conservative | 44.63 (42.27-47.49) | 12.2% (9.8%-14.2%) | | 14,010 (6,516-28,606) | 12,050 (5,700-23,394) | | 2.26 (1.56-3.27) | 13.2% (10.0%-18.8%) | | 18.86 (16.27-22.43) | 2.1% (1.4%-3.1%) |
| Optimistic | 52.83 (50.44-55.35) | 32.8% (27.9%-36.3%) | | 20,272 (10,455-38,496) | 17,154 (9,249-31,212) | | 1.96 (1.37-2.81) | 24.6% (20.1%-30.0%) | | 18.50 (16.02-21.92) | 3.9% (2.8%-5.2%) |

*We sampled for 58 parameters for the sensitivity analysis, including key PrEP-related parameters (PrEP effectiveness, reduction in condom use while on PrEP, tail protection duration) and the cost of service provision of LEN and CAB, and sampled from pre-determined distributions for each of the 1,000 model runs (see Table S1). Abbreviations: HIV=Human immunodeficiency virus, AIDS = acquired immunodeficiency syndrome, CAB-LA = long-acting injectable cabotegravir, USD = United States Dollars, BL = Baseline, PrEP = pre-exposure prophylaxis

1. Johnson L, Dorrington R. Thembisa version 4.8: a model for evaluating the impact of HIV/AIDS in South Africa. 2025 https://thembisa.org/content/downloadPage/Thembisa4_8report (accessed April 14, 2025)
